# Exploring virtual and augmented reality simulation in obstetrics and gynaecology training; a qualitative study

**DOI:** 10.1101/2020.12.13.20248133

**Authors:** R Ryan, G Ryan, MF Higgins

## Abstract

**Introduction:** The scope and use of simulation in medical education and healthcare training has increased considerably over the past 20 years, as technology has advanced rapidly and increasing fidelity has become standard.

This study aims to explore O&G trainees’ attitudes and beliefs regarding augmented reality (AR) and virtual reality (VR), and their attitudes towards their incorporation into relevant curricula.

**Methods:** This was a qualitative descriptive study using thematic analysis. An initial pilot focus group was followed by semi-structured interviews. Data was analysed using iterative content analysis. Relevant themes were identified during the focus ground and explored in greater detail during semi-structured interviews.

**Results:** Nine trainees participated in total; one focus group and two individual interviews were conducted. Five key themes were identified: the value and importance of education, ‘what works’, the importance of a supportive educator and peer, the difference between simulation and real-life, and VR-associated hope.

**Conclusions:** Trainees had little experience or knowledge of AR and VR simulation. They believe that simulation will be more important in future, and that it has potential for training. Further exposure and education involving AR and VR will be required for it to become an integral part of education and training.

## Introduction

Simulation is defined as ‘*the technique of imitating the behaviour of some situation or process (whether economic, military, mechanical, etc*.*) by means of a suitably analogous situation or apparatus, especially for the purpose of study or personnel training*.’ (OED 2006, in Bradley 2006). In medical education, the use of virtual reality and augmented reality simulation is increasing in popularity. Virtual reality refers to the recreation of environments or objects as a complex, computer generated image. Augmented reality refers to the overlaying of a computer-generated image on a real-life scenario. Haptic systems refer to those replicating the kinesthetic and tactile perception and are often used in conjunction with both virtual and augmented reality systems in order to increase fidelity. (Bradley 2006). Simulated training is increasingly being seen as a moral imperative. Following the publication of ‘*To Err is Human’*, (Kohn 2000) it is increasingly recognized that patient safety is all too often not the core focus of healthcare systems. Patient care must now, rightfully, take priority, with medical education requiring a shift in its delivery. Simulation has become a viable alternative for early training and skill acquisition, allowing learners to gain familiarity and competence with tasks and situations prior to direct patient contact.

There are many advantages of simulation in obstetrics and gynecology (O&G) postgraduate education. It allows learners to repetitively practice tasks, procedures and skills in order to improve performance. Furthermore, tasks/scenarios can be created on demand and tailored to individual learners’ skills (Bradley 2006). The scope and use of simulation in medical education and healthcare training has increased considerably over the past 20 years, as technology has advanced rapidly and increasing fidelity has become standard. Simulation is increasingly being used in O&G worldwide. The American College of Obstetrics and Gynecology have established a simulation working group in order to construct a simulation curriculum encompassing many common procedures and situations encountered in the field (ACOG 2018).

Worldwide, there is increasing interest in trainee’s experiences of simulation across a variety of medical and surgical fields. An evaluation of surgical resident’s perceptions of simulation as a clinical learning approach (which did not specifically evaluate resident’s perceptions of virtual reality and augmented reality simulation), noted that trainees had a very narrow perception of the utility of simulation (Walsh et al. 2017). They viewed it largely as an introduction to task-based learning rather than as a tool for ongoing learning, or for skills other than ‘hard’ skills such as surgical techniques. A qualitative analysis of medical student’s experiences of simulator-based teaching, found that 94% students found simulation to be excellent, whilst valuing its utility in thematic domains from knowledge to skill acquisition to teamwork and communication (Takayesu et al. 2006). In obstetrics specifically, a qualitative study explored Brazilian O&G resident’s perception of a postpartum hemorrhage simulation training and found that all residents interviewed felt that they achieved long term transfer of learning following simulation (De Melo 2018).

To the authors’ knowledge, there have been no formal studies to date investigating trainee’s knowledge, beliefs and attitudes surrounding AR and VR simulation.

The importance of user-centered development in AR has been previously highlighted, and an understanding of learner’s needs and perceptions is required to ensure the development of appropriate materials (Thomas 2010). This research would provide valuable insights into future priorities for residency curricula, and opportunities for expanding the scope of virtual and augmented reality simulation in specialties such as O&G.

The aims of this research project were therefore to explore O&G NCHDs’ attitudes and beliefs surrounding AR and VR and to explore O&G NCHDs’ attitudes toward the incorporation of AR and VR into the relevant curriculum. The hypothesis was that qualitative research would provide a rich approach in which to fully explore themes.

## Methodology

This study took place amongst non-consultant hospital doctors (NCHDs) working in a tertiary maternity hospital. The study comprised of both doctors enrolled in the formal Irish O&G training scheme, and doctors undertaking 6-month non-training posts. The Irish O&G training scheme is an eight-year training scheme with a formal curriculum, with elements of AR and VR training available to trainees in a variable way. There is no standardized AR or VR training component. The hospital site involved does not currently have AR or VR training available on site; this is being developed at present as part of this research project.

Qualitative methodology, with data presented in the everyday language of the participants and with projects informed by prior expert knowledge (Neergard 2009) was used in this research project. An initial pilot focus group, followed by semi-structured interviews, comprised the research study. Study participants were selected by purposive sampling from a sampling frame of non-consultant hospital doctors in the study institution. Maximal variation sampling was used (Robinson 1999; Kahlke 2014). Aside from employment, participants varied in level of experience, ethnicity, gender, training scheme status and age.

Participants were invited to participate by an email invitation. There was also an advertisement posted in the doctor’s room of the study institution with information regarding the research study and a contact email for those interested in participating. The relevant gatekeeper (the assistant master of the hospital), was approached and agreed to facilitate recruitment.

Informed consent was obtained from all participants prior to study participation. With regard to confidentiality and anonymity, prior to commencement of the focus group, participants were advised that all information shared during the focus group should remain confidential and not be discussed outside of the focus group. Collected data was coded and was not shared outside of the research group (including with participants) prior to publication. Every effort was made to anonymise responses, however given the small sample size, participants retained the right to request removal of information that might allow participants to be identified, from written transcripts.

Sample size was determined when data saturation was reached during semi-structured interview, suggesting no new information is forthcoming (Tavakol and Sandars 2014).

Data collected was audio recorded and transcribed by the primary researcher (RMR). The data was re-examined after transcription by the three study investigators (RMR, GR, MFH). Study participants were given the opportunity to review the transcripts and make any changes they considered necessary.

Data from the initial focus group was analysed in an iterative manner, i.e. was conducted at the same time as data collection, in order to guide further sampling for semi-structured interviews, identify emerging findings, and identify when no new information was apparent (data saturation). Content analysis was chosen in order to depict the rich and detailed information provided by study participants (Hsieh and Shannon 2005). Inductive content analysis was used in order to identify core themes. Coding and identification of themes was carried out independently by the three researchers, following which a discussion and merging of relevant themes formed the basis of the final themes identified.

Core themes previously identified during the pilot study were further explored during semi-structured interviews. Data from the semi structured interviews was analysed using deductive content analysis, where previously identified themes and the study participant’s reaction and responses to those themes were identified, isolated, and then aggregated with the responses of other study participant’s regarding that same theme. Once again, identification of further themes raised during the semi-structured interviews was carried out by two researchers independently (RMR, MH), and the final themes were a composite of the themes that both researchers identified. These themes that were developed were therefore prevalent throughout interviews, important and substantial. Thematic saturation was defined when refinements did not add substantial new themes.

Trustworthiness was enhanced by two methods, member checking and triangulation. Member checking was performed by both checking themes identified with the original participants and with the individuals during semi-structured interviews. Triangulation was based on use of two initial researchers developing themes (RR. MH) and a third reviewing these themes (GR).

Data analysis software (N-Vivo10; QSR International 2012) was used in the thematic analysis of the pilot focus group. In coding, participants in the focus group study were labelled ‘P’ followed by A-G. Participants in the semi structured interviews were labelled ‘OO’ followed by a digit 1-2. Research interviews occurred during the academic year September 2018-July 2019.

Members of the research team had reflected a priori on possible personal sources of bias: all three are enthusiastic medical educators with an interest in innovation and research.

Ethical approval was obtained from the relevant hospital and university ethical approval committees. This study is reported using the SRQR standards (O’Brien et al. 2014).

## Results

One focus group and two individual interviews were conducted with a total of nine NCHDs. All participants were working in O&G in Ireland. Table 1 demonstrates the participant characteristics. Seven participants were female. Four participants were on the higher specialist training scheme (years five to nine following graduation from medical school), three participants were on the basic specialist training scheme (years two to four following graduation) and two participants were undertaking non training posts.

**Table 1.** Participant Characteristics

| Participant characteristic | N (%) |
| --- | --- |
| Female | 7 (78) |
| Irish trained | 7 (78) |
| On a training scheme | 7 (78) |

Five overarching themes were identified during the pilot focus group discussion, and further clarified during semi-structured interviews. Table 2 displays these overarching themes and the relevant associated subthemes.

**Table 2.** Identified themes and associated subthemes

|  |  |
| --- | --- |
| Value and importance of education | • Respect for the importance of ‘knowing’ for clinical care |
| What works | • The value of the visual<br>• Recognizing different needs for different procedures<br>• Significance of having easy access to educational tools.<br>• Importance of real-life in building on experience, non-technical skills and teamwork |
| Importance of a supportive educator / peer | • Supportive and experienced Educator<br>• Peer to peer learning |
| Difference between simulation and real-life | • Additional complexity of real life<br>• Training versus reality |
| Augmented Reality raised concerns; Virtual reality raised hope | • Concern for the future<br>• Wary of dependency on technology |

### Value and importance of education

#### Respect for the importance of knowing for clinical care

Trainees showed great respect for the importance of getting a correct diagnosis or performing a procedure correctly – “..*missing ectopics…quite dangerous*” [PF] as well as a wish to have more confidence in making a diagnosis when it was correct “..*pick up findings I am afraid of mentioning…need someone to come and make sure you picked up the right thing*” [PE]. It was important not to succumb to pressure to scan a patient and discharge her home, rather than waiting for back up to confirm or refute a presumed diagnosis – “…*if I could just do this then the patient could go home, but I think it’s safer not to be told to just go and do this scan*..” [PC].

These findings were echoed in the semi-structured interviews “*you can learn the anatomy in a book, but have to be doing diagnostic [laparoscopy] to know the anatomy in order to be able to do it on your own*”[OO2], and “*Ultimately I think you shouldn’t need the prompts to do it, you should know*.” [OO1]

### What works

#### The value of the visual

Many of the trainees in the focus group used ‘YouTube’ videos in preference to books, as they allowed visualisation of the procedure in real time. Books were used but sometimes it was “*difficult to visualise what they are describing*” [PC] or were not as realistic “*no blood in the books”* [PC] and “*everything is colour coded*” [PF]. Real-life experience trumped both books and videos “*it’s important to supplement with something but it won’t replace it* [PG]”

Exploring this theme further in semi-structured interviews, trainees again reiterated their preference for YouTube videos, noting its utility and convenience - “*when it’s on a screen you can watch it pretty much anywhere*” [OO2]. One respondent reported that he preferred YouTube videos to watching something in real life -”*live demonstration is good, but I think I prefer demonstration on a screen because you can see everything more clearly*”[OO2]. This contrasted with the responses of the focus group and the other semi-structured interviewee “*I’d get most from watching something being done procedurally*” [OO1].

#### Recognising different needs for different procedures

Trainees described the different senses used for different procedures – “blind” procedures (such as the surgical procedure for evacuation of retained products) where touch was the most important sense for feedback, “*you’re dependent on your own feel and experience*” [PA] compared to more visual open procedures “*you can see what you’re doing, it’s different, different story*” [PA].

#### Significance of having easy access to educational tools

Trainees clearly recognised the value of prior learning prior to working in real-life situations “…*to prepare you because you need to have some form of skill. You can’t practice on real people*” [PF]. While trainees have access to a simulator for trans-vaginal ultrasound “*it’s incredibly difficult to access them. With work and we don’t get time*” [PA]. As higher trainee’s they have access to a simulator that they can use at home “*it’s great that you actually get it*” [PB] “like a whole month” [PA] but still no substitution for having a trainer as well “…*someone training you and you have support*” [PG]

This theme was emphasized in semi-structured interviews, with trainees reporting “*First of all you need to have a place that is readily accessible to do [training], and secondly you need to have the time to do it*.” [OO2] Trainees commented on the difficulties of availing of AR and VR simulation training within the training system, commenting that barriers were “*…access mostly, it’s [ultrasound simulator] locked away, you have to email ahead* [to get access], *it is really only available office hours, even if you’re free Saturday or Sunday it’s not available*.”

#### Importance of real-life in building on experience, non-technical skills and teamwork

Trainees recognised the importance of experience to underpin both theory and practical education – the value of doing a procedure so many times that they build up a muscle memory “*that is just through doing. ‘Cos it’s the manipulation and your orientation, knowing where your hand is compared to what you see on the screen which I think is just a practice thing*” [PC]. Working in teams in real-life was different to learning as an individual “*lots of people in the room it’s different*” [PG]

Semi-structured interviewees’ views chimed with those of the focus group in this regard. “*doing the real thing is a whole different thing. Getting it done instead of as practice…because you’re in the atmosphere*.”[OO2] “*The OR is just more tense; different dealing with a live subject, time is different you can’t take an hour to do a diagnostic laparoscopy but you could spend as much time as you want with the simulator*”[OO1]. These views were shared by respondents of varying experience levels.

### Importance of supportive teacher / peer

#### Supportive and experienced teacher

Having a supportive and experienced educator was a constant theme identified as being invaluable. Such an educator could provide the theory to underpin the practice *(“no one had ever explained that to me before…there’s probably more that we could get from being taught*” [PB]) as well as providing feedback when procedures did not work out as planned “*when you’re wrong you can’t really figure out why you’re wrong…you still need a teacher”* [PG].

Semi-structured interviewees shared the view that a mentor was an invaluable asset “*If you enjoy learning off someone you get motivated to learn from them, and you get motivated to learn as well. I really think that having a mentor figure is huge, and I don’t think that only applies to medicine I think that’s everywhere*.” [OO1]

One respondent compared a supportive and experienced educator to a guide “*I don’t really think you’re practicing on patients because the consultant with you is doing the operation through your hands*” [OO1]

#### Peer-learning

Peer to peer learning was also identified as a common method of learning on-the job - “*I learned it on the go, as a colleague of mine, another SHO* [year 2-3 post graduate doctor], *told me, you learn one you do one you teach one right*?” [OO2].

Dyad learning (learning in pairs with support from a facilitator) was also useful “*there was someone else there, that built-in peer trainer*” [PG] not only as it made trainees “*always more inclined to do something if you can have a friend*” [PB] but it also “*made it competitive*” [PC].

#### Difference between simulation and real-life

When comparing transvaginal ultrasound using a simulator versus a real-life patient there were clear differences “*don’t actually think the movement is anyway the same*” [PG]. Real-life was the true training “*real-life, doesn’t replace it*” [PC] “*can’t mimic it*” [PA] “*you don’t have the same pressure*” [PC] though simulation was a “*good step and helpful to do*” [PF]. Clear advantages were expressed on first learning on a simulator with support (“*good theoretical plus practical training and then let people practice*” [PF]) as they gave confidence (“*oh yeah, I can do this*” [PB]), and provided a muscle memory (“..*a bit slicker with movement*” [PA], “*comfort”*[PB]) and provided *“familiarity with instruments*” [PA], helped “*hand eye coordination*” [PF] “*depth of perception*” [PA] as well as having a safe environment to learn “*I wonder what happens if I do thi*s” [PB].

This theme was elaborated upon during semi-structured interviews, with one trainee making the point that there was value in teaching people how to fail “*Doing this AR there should be a massive leeway in being allowed to fail. Because that’s pretty much the best way we learn – to find out where we went wrong*.”, “*you learn more from failing then from doing something right you know? When you do something right, you’ve done it right but there could have been some things that could have been done better and these things I think we only learn after we fail or make mistakes*” [OO2]

#### The concerns of Augmented Reality and the Hope for Virtual reality

Most of the trainees had little exposure to augmented reality; those that did were through entertainment (“*Disneyland*” [PA], “*walk through a jungle*”, “*on a roller-coaster*” [PG]) rather than educational. Concerns were raised about the validity of AR in education, whether it would be “*distracting*” [PF], how they would deal with “*anatomical variations*” [PC] and concerns that teaching supports would be withdrawn “*people won’t teach you anymore*” [PG] These concerns were shared by trainees during the semi-structured interviews, but to a lesser degree. There was an acknowledgement that this method of learning will be more important in the future “*Because everyone’s anatomy is a little different, so it has to be tied to individuals’ anatomy. I guess I don’t think it’s there yet, but I definitely think that’s where it’s going*.” [OO1]

Virtual reality in contrast was a possible learning tool for both non-technical skills and teamwork, where VR “*would be helpful*” [PC]. There was a clear difference between learning an emergency as an individual “*you can list*..*what to do*” compared to the teamwork required “*there’s lots of people in the room its different. Learning that is harder*” [PG].

## Discussion

This study, to our knowledge the first to examine O&G trainee experiences, knowledge and beliefs surrounding AR and VR, suggests trainee’s experience of AR and VR is limited. Many trainees report video games and theme parks as their sole exposure to AR and VR. Within a work environment, multiple barriers to accessing simulation training were identified, including time, availability, accessibility and geographic restrictions. VR in particular was viewed positively, with trainees viewing it as a useful adjunct. However, a dominant message throughout the study was that AR and VR could never be used to replace real-life experiences.

A qualitative study design was appropriate to investigate the research question as the aim was to understand the beliefs and attitudes of O&G trainees and gain an in-depth understanding of their attitudes, rather than quantify the use of this technology. Initially, an ‘emic’ or inductive approach to the topic was used, as during the pilot focus group interview researchers attempted to put aside their prior theories and assumptions regarding participants’ knowledge, beliefs and attitudes towards AR and VR in O&G, and allow themes, patterns and concepts to crystallise during the focus group process. This involved an element of reflexivity on the researcher’s part, particularly as the study participants were co-workers and peers. Following this initial exploratory focus group, semi-structured individual interviews were carried out with key informants using an ‘etic’ or deductive approach; contrasting the theories identified from a search of the literature with themes that emerged from the focus group interview. This further elucidated the beliefs and attitudes of the study population and placed them within the context of wider research on this topic.

Studies conducted amongst other surgical specialties found that surgical residents echoed the view of the trainees in our study. Walsh et al (2017) evaluated surgical resident’s perceptions of simulation as a clinical learning approach. While they did not specifically evaluate resident’s perceptions of virtual reality and augmented reality simulation, they note that trainees had a very narrow perception of the utility of simulation, and viewed it largely as an introduction to task based learning rather than as a tool for ongoing learning, or for skills other than ‘hard’ skills such as surgical techniques. These views could be seen as at odds with the limited published data on the utility of AR and VR in laparoscopic and hysteroscopic O&G training. Larsen et al (2009) used LapSimGyn in a Danish cohort of O&G trainees (SHO equivalent) and found that 8 hours of simulation training was equivalent to performing 20-50 laparoscopic cases compared to controls. Virtual reality laparoscopic tubal ligation simulation has also been validated for salpingectomy (Johnston et al. 2015). Alici et al (2014) carried out a prospective cohort study which demonstrated that training on a hysteroscopic augmented reality simulator with haptic sensors (EVA ETXY/Hystero; Prodelphus, Brazil) improved medical student’s skills as measured by an objective structured assessment of technical skill (OSAT) as well as self-assessed skills. Elessawy et al (2017) used the virtual reality HystSim trainer to evaluate the efficacy of the trainer to improve the acquisition of hysteroscopic skills in both novice and experienced hysteroscopists.

Strengths of this study include the intensive study of the small number of participants, ensuring adequate breadth and depth of information on a topic that participants had not had much exposure to previously. Participants were chosen purposefully, with selection being sequential as the interview material evolved (Curtis et al. 2000). This ensured rich data collection, which is exemplified in the datasets congruent themes which are well developed and explored thoroughly during this analysis.

One weakness of this study is the potential bias introduced at various stages of the research process. This study is an example of practitioner research which has many advantages in a qualitative research capacity as the researcher has prior knowledge and experience in the field. However, this approach can introduce bias, including reporting bias, with participants unwilling to discuss negative experiences with simulation during the focus groups (Smyth 2008).

In order to minimise bias in data collection, two approaches were used. The research team reflected a priori on their possible biases in the research area and the primary researcher maintained a reflexive diary, in order to continually reiterate the core focus of the inductive approach to the focus group (DePoy 2010). During the semi-structured interviews, techniques such as active listening, open body language, appropriate use of silence, echoing of participant’s language, and linking phrases which encourage participants to discuss their comments in more detail were used, both to improve participant’s responses but also to minimise researcher input that may introduce bias (Creswell 1998). During data analysis and interpretation, peer debriefing was conducted with a research supervisor who had not been involved in data collection, a technique shown to improve data validity (Smyth 2008).

Another potential weakness of this study is the low number of participants involved. Despite the low study numbers, data saturation was reached within the study, with themes reiterated from the focus group in semi-structured interviews, no new codes emerging, and given the theory, enough data to illustrate the points made (Saunders et al. 2018; Starks and Trinidad. 2007). Views were, in general, consistent amongst trainees.

This qualitative study provides an in-depth exploration of trainees in O&G in one hospital in Ireland’s experiences of AR and VR simulation. In order to further this research, a mixed methods approach could be employed. Based on the findings of this research, a quantitative questionnaire could be developed specifically asking about themes raised during this study: knowledge of AR and VR simulation; access to AR and VR simulation; perceived utility of AR and VR simulation. This could be distributed to a wider network of trainees in O&G, in order to expand the scope of the research.

In conclusion, this study identifies existing knowledge surround AR and VR amongst O&G trainees, explores a variety of beliefs surrounding its potential as an educational tool, and places it within a context of workplace-based learning tools.

### Practice points

|  |
| --- |
| 1. Simulation is becoming increasingly prevalent in O&G training, both at an undergraduate and post-graduate level, and will only increase in importance following the COVID-19 pandemic |
| 2. Trainees recognize many potential benefits of AR and VR, but have limited exposure to simulation as training |
| 3. Simulation in conjunction with expert or peer to peer learning was seen as superior to simulation alone for surgical learning in O&G |
| 4. Simulation is currently viewed as a useful ‘first step’ and adjunct to real-life training, but is not viewed as equivalent to real-life training |
| 5. Further exposure will be required to convince trainees of the validity of simulation in training |

## Data Availability

Data is available from the authors under reasonable application

## Notes on contributors

Dr Roisin Ryan is an OBGYN trainee who has worked in Ireland, the UK and the USA. She is currently completing residency in OBGYN at the Brigham and Women’s Hospital and Massachusetts General Hospital, Boston. She holds a Master’s in Education, and her research interests include technology in clinical education.

Dr Grace Ryan ***

Dr Mary Higgins ***

